# From loss to recovery: how to effectively assess chemosensory impairments during COVID-19 pandemic

**DOI:** 10.1101/2021.03.25.21254253

**Authors:** Cinzia Cecchetto, Antonella Di Pizio, Federica Genovese, Orietta Calcinoni, Alberto Macchi, Andreas Dunkel, Kathrin Ohla, Sara Spinelli, Michael C. Farruggia, Paule V. Joseph, Anna Menini, Elena Cantone, Caterina Dinnella, Maria Paola Cecchini, Anna D’Errico, Carla Mucignat-Caretta, Valentina Parma, Michele Dibattista

**Affiliations:** Department of General Psychology, University of Padova, Italy; Leibniz-Institute for Food Systems Biology at the Technical University of Munich, Germany; Department of Physiology, Monell Chemical Senses Center, USA; Private practice VMPCT, Milan, Italy; ENT department, Italian Academy Of Rhinology - ASST sette laghi Varese; Institute of Psychology, University of Muenster, Germany; Department of Agriculture, Food, Environment and Forestry (DAGRI), University of Florence, Italy; Interdepartmental Neuroscience Program, Yale University, USA; National Institutes of Nursing Research; National Institute of Alcohol Abuse and Alcoholism; National Institutes of Health; Neurobiology Section, SISSA, International School for Advanced Studies, Italy; Department of Neuroscience, ENT section, Federico II University of Naples, Italy; Department of Neurosciences, Biomedicine and Movement Sciences, Anatomy and Histology Section, University of Verona, Italy; Department of Neurobiology, Goethe Universität Frankf urt, Germany; Department of Molecular Medicine, University of Padova, Italy; Department of Psychology, Temple University, USA; Department of Basic Medical Sciences, University of Bari A. Moro

## Abstract

Chemosensory impairments have been established as a specific indicator of COVID-19. They affect most patients and may persist long past the resolution of respiratory symptoms, representing an unprecedented medical challenge. Since the SARS-CoV-2 pandemic started, we now know much more about smell, taste, and chemesthesis loss associated with COVID-19. However, the temporal dynamics and characteristics of recovery are still unknown. Here, capitalizing on data from the Global Consortium for Chemosensory Research (GCCR) crowdsourced survey, we assessed chemosensory abilities after the resolution of respiratory symptoms in participants diagnosed with COVID-19 during the first wave of the pandemic in Italy. This analysis led to the identification of two patterns of chemosensory recovery, limited (partial) and substantial, which were found to be associated with differential age, degrees of chemosensory loss, and regional patterns. Uncovering the self-reported phenomenology of recovery from smell, taste, and chemesthetic disorders is the first, yet essential step, to provide healthcare professionals with the tools to take purposeful and targeted action to address chemosensory disorders and its severe discomfort.

## Introduction

According to the World Health Organization, COVID-19 has been confirmed in more than 113 million cases across 223 countries, leading to more than 2.5 million deaths (https://www.who.int/emergencies/diseases/novel-coronavirus-2019, Last update: 27 February 2021). Recent estimates indicate that up to 98% of individuals diagnosed with COVID-19 developed forms of chemosensory disorders, most prominently smell loss^1–7^. Data collected before the COVID-19 pandemic showed that up to 49% of the population report an episode of olfactory loss over their lifetime, with 5% of them reporting complete smell loss (anosmia)^8–10^. Population-based epidemiological studies before COVID-19, mostly focused on older adults^10–15^, provide prevalence estimates of smell loss ranging from 2.7% to 24.5%^16–20^ and taste disorders ranging from 0.6% to 20%^11,18,20–22^. Reports to date reveal that the COVID-19 pandemic has already significantly increased the prevalence of chemosensory disorders worldwide, especially among younger cohorts^1,23^, yet the global estimates on chemosensory disorders may be markedly underestimated.

Chemosensory disorders are both early and specific symptoms of COVID-19^24–26^. Previous studies indicated that the timeframe for a full or partial recovery (in particular of the sense of smell) seems to be highly variable, spanning from 8 days to even 8 weeks^3,27–32^. For the vast majority of patients (up to 85%), chemosensory issues resolve along with Covid-Like-Illness (CLI) symptoms, in approximately three weeks^31–35^. Nevertheless, approximately 15% of patients continue to report chemosensory loss as their main neurological sequela, which persists after the resolution of CLI symptoms^32,33^. Therefore, if patients affected by COVID-19 are initially very concerned about the development of the infection and the severity of the illness, in a later stage, they develop serious concerns for a prompt resolution of smell and taste loss. Persistent smell and taste loss are an unexpected and invisible disorders associated with a significant reduction in a person’s quality of life^36–38^, including increased depressive symptoms^36^, anxiety^39^, sexual desire^40^, nutritional^41–44^, and safety issues^36,45^. It is important to note that these side effects are not COVID-specific but characterize patients’ experience affected by smell and taste loss because of a variety of etiologies^36,45^. Unexpectedly, during the COVID-19 pandemic, smell and taste loss took center stage, likely due to a reduced awareness of chemosensory disturbances by clinicians and the public^46^. As a result, national healthcare systems worldwide might often not be well prepared to address the needs of patients who suffer from smell loss long-term.

Non-profit associations have emerged to fill this gap and support patients in their journey to gather information on their condition, its consequences, and provide validation of this invisible sensory experience and emotional support (https://gcchemosensr.org/patient-orgs/). Italy has been the first European country to be massively hit by COVID-19^47,48^ (http://www.salute.gov.it/imgs/C_17_notizie_4403_0_file.pdf), the Lombardy region was particularly affected, reaching the highest death toll of the first wave (28K on 25 February, 2021; https://www.statista.com/statistics/1099389/coronavirus-deaths-by-region-in-italy/)^49^. As a result, throughout the national territory, and in particular in the most affected regions, the need to address COVID-19 long-haulers with chemosensory symptomatology has emerged early and prominently^50–53^. The Italian National Healthcare system currently lacks capillary specialized assistance for patients with smell and taste loss. Approximately 5500 otolaryngologists operate in the country, of which only a minority is specialized (approximately 5% of them) in taste and smell disorders (interview with Carmelo Zappone, president of Associazione Italiana ORL Libero Professionisti, https://www.aiolp.it/). Several initiatives are currently being developed to face the new challenge, e.g., 31 ENT centers affiliated with the Italian Academy of Rhinology are currently offering training courses and sniffing test kits (e.g., Sniffin’Sticks test^54^) to identify and monitor patients with smell loss (https://accademiarinologia.it/index.php/it/). Taste and smell specialists are mostly located in clinics and centers within hospitals, yet the emergency measures undertaken in response to the COVID-19 pandemic have drastically reduced ENT outpatient activities^55,56^.

Therefore, to face the exponential increment of patients with taste and smell disorders, a greater number of healthcare professionals, including general practitioners as well as frontline healthcare workers, would need tools to recognize and validate the subjective chemosensory experience of patients to refer them to specialists.

No routine chemosensory testing is available to date for healthcare professionals to be used on a large scale, therefore studying the phenomenology of recovery from chemosensory loss, including smell, taste and chemesthetic manifestations based on patients’ self-reports is the first, yet essential step, to provide the tools to take purposeful and targeted action to address chemosensory loss and its significant discomfort in patient’s lives. Here, we tested our pre-registered hypotheses on the self-reports of the Italian participants collected via the crowdsourced GCCR survey^57^ detailing the phenomenology of self-reported chemosensory abilities before, during and after COVID-19 diagnosis. First, we aimed to describe the patterns of recovery of smell, taste, and chemesthetic abilities, individually and in combination, in relation to the timeline of other CLI symptoms. We set out to confirm that the chemosensory recovery would be more advanced the farthest from CLI symptom onset and for limited losses during the disease; we explored the synergies between chemosensory modalities in promoting successful chemosensory recovery based on their profiles of loss during the disease. Second, we assessed whether specific demographic information, COVID-19 symptoms and/or prior medical conditions constitute risk factors for lengthy or no recovery from chemosensory loss within 6 months.

### Partial or substantial chemosensory recovery from COVID-19

Data from a final sample of the 974 Italian residents who participated in the GCCR online survey between 10th of April 2020 and 17th of October 2020 and who reported partial or full recovery from CLI was used to determine profiles of recovery patterns (Figure S1 for details on the full sample). With the goal of limiting the number of questions that a healthcare professional should ask to determine the state of chemosensory recovery, we focused on rating scales, which proved to be the most predictive way to identify individuals positive for COVID - 19^24^. We therefore selected the ratings on 0-100 scales given to smell, taste and chemesthesis abilities (i.e., the ability to perceive the spiciness of chili peppers, the cooling of menthol and the carbonation in soda) after the disease minus their ratings during the disease. Participants reported to have significantly lost their sense of smell (mean = 11.90, SD = 23.47), taste (mean = 20.39, SD = 28.07) and chemesthesis (mean = 40.81, SD = 32.97) during COVID-19 as compared to before COVID-19 started [smell: mean = 91.14, SD = 16.82 t(973) = 82.71, p < 0.001; taste: mean = 92.74, SD = 13.71 t(973) = 70.14, p < 0.001; chemesthesis: mean = 87.63, SD = 17.30 t(973) = 38.99, p < 0.001, Figure 1A]. After the resolution of CLI symptoms (Figure 1A), on average smell (mean = 53.05, SD = 32.22; t(973) = 35.98, p < 0.001), taste (mean = 60.75, SD = 30.89; t(973) = 35.39, p < 0.001) and chemesthesis abilities (mean = 69.52, SD = 25.80; t(25.69) = 973, p < 0.001) improved. However, such post-CLI improvement is not homogenous. An exploratory cluster analysis (k-means, bootstrapped stability = 0.98) revealed two chemosensory recovery groups: partial (N=471, 48.36% of the sample; centroids: smell = 13.4, taste = 10.07, chemesthesis= 7.55) and substantial (N=503, 51.64% of the sample; centroids: smell = 67.2, taste = 68.1, chemesthesis = 48.5; Figure 2A, B). The three chemosensory modalities contributed equally to Dimension 1 that explained 73.2% of the variance while chemesthesis recovery was the major contributor to Dimension 2 (Figure 2B).

**Figure 1.**
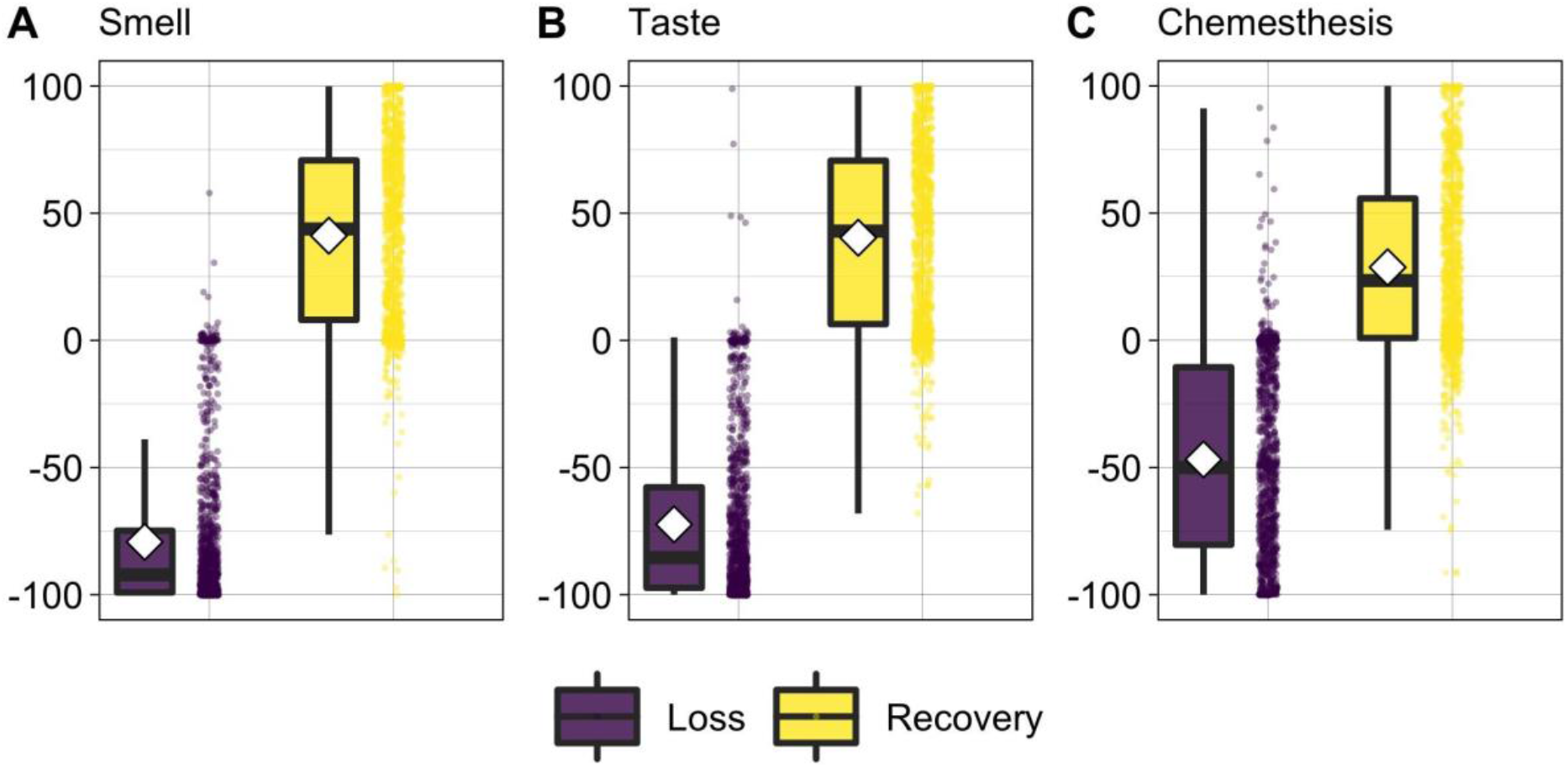
Flow diagram presenting the selection of the observations included in the present study.

**Figure 1.**
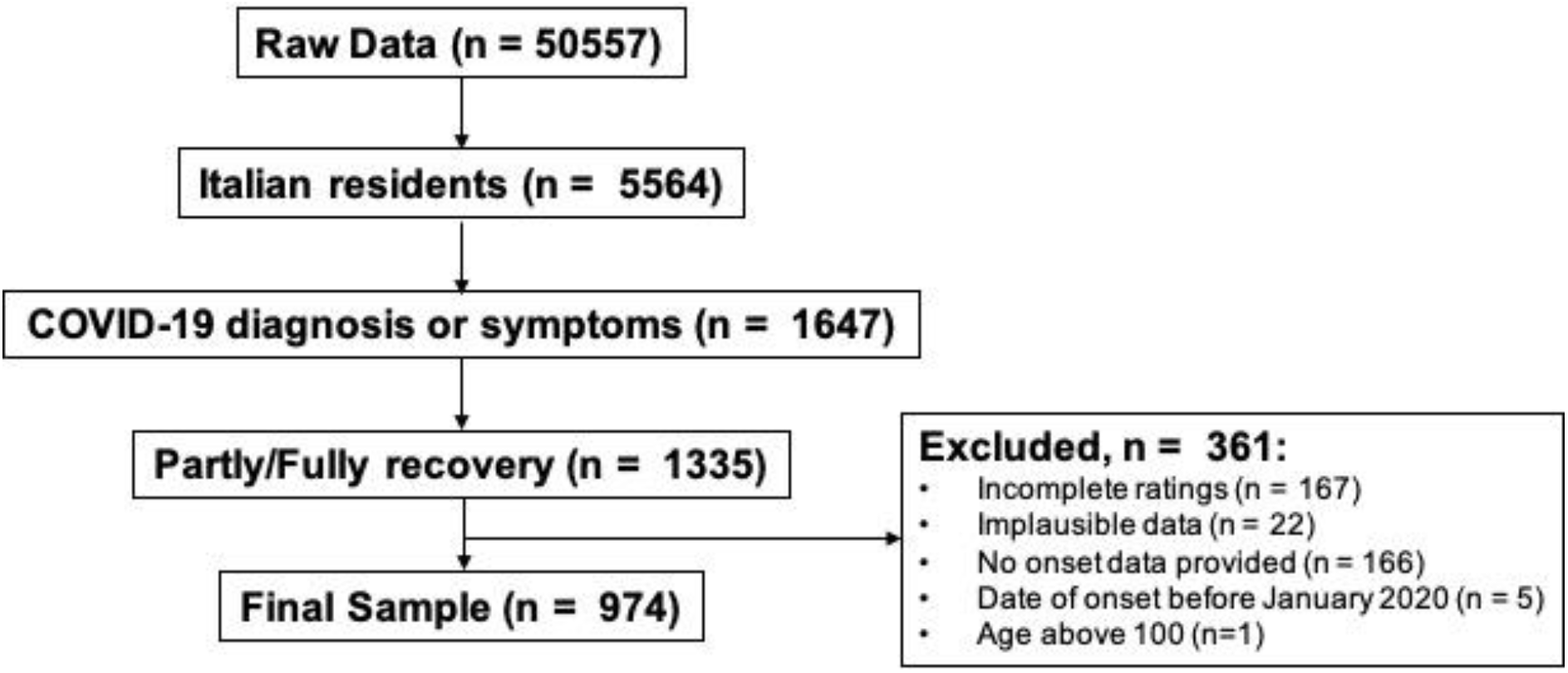
Loss (during - before ratings; violet) and recovery (after - during ratings; yellow) of smell (A), taste (B), and chemesthesis (C). Boxplots depict the median (horizontal black line) and quartile ranges of the distribution; white diamonds indicate the mean; whiskers indicate maximum and minimum values. The raw data are shown as dots to the right of each boxplot.

**Figure 2.**
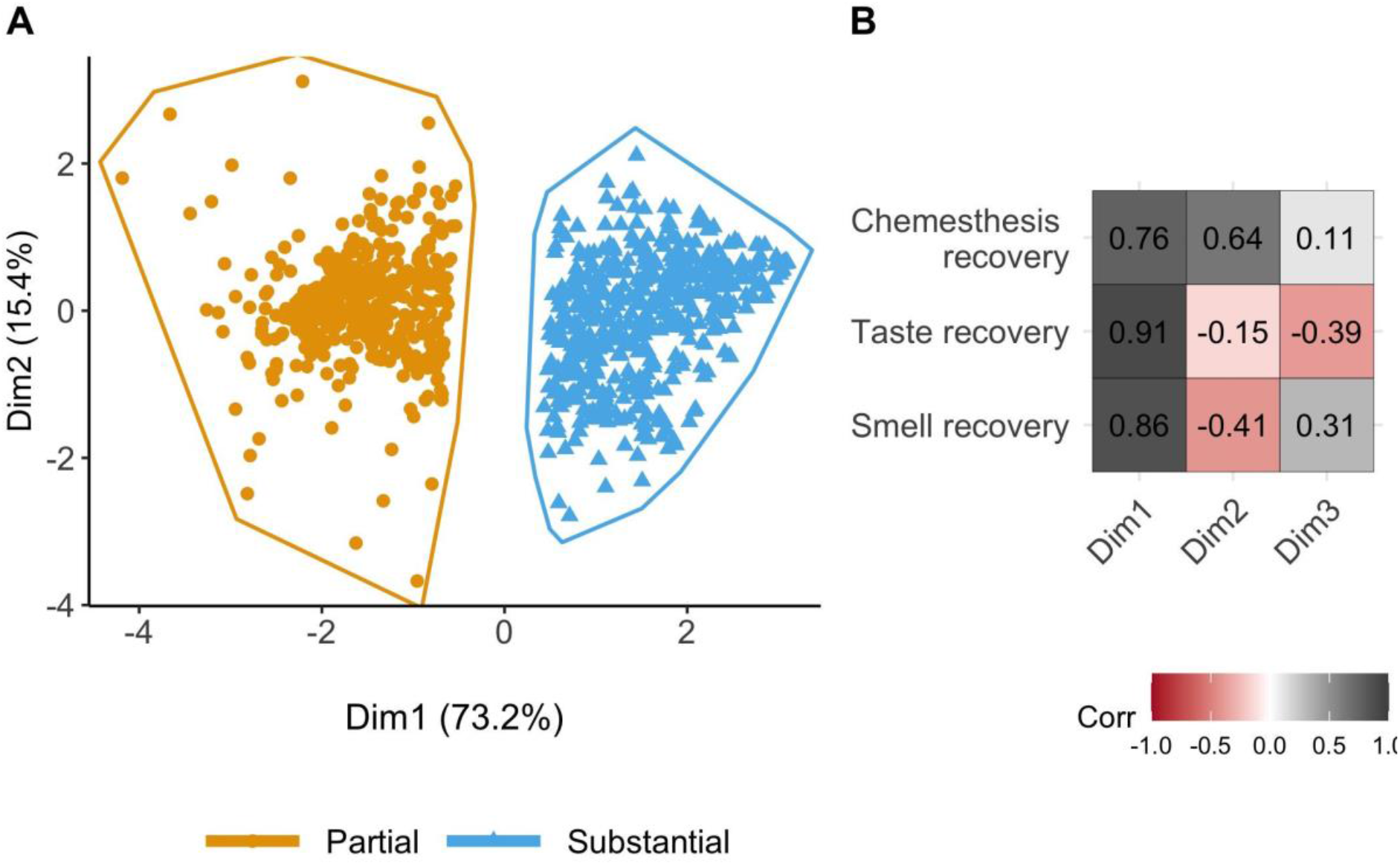
A) Clusters of participants on chemosensory recovery identified by *k-means* clustering. The scatterplot shows each participant’s loading on Dimension 1 (Dim1) and Dimension 2 (Dim2) of the Principal Component Analysis. Partial = smell, taste, and chemesthesis partial recovery; Substantial = smell, taste, and chemesthesis substantial recovery B) Correlations between the three principal components with respect to recovery in smell, taste, and chemesthesis. Gray color indicates a positive correlation, whereas shades of red indicate negative correlations. Darker shades indicate stronger correlations.

Among other characteristics (Table 1), participants who only partially recovered their chemosensory abilities at the time of survey completion were older (mean = 43.16, SD = 11.74) and reported to have contracted the disease earlier (mean = 43.15 days, SD = 23.87) than those who substantially recovered [age: mean = 39.63, SD = 10.75, t(949.86) = 4.88, p < 0.001; days from COVID-19 symptom onset: mean = 40.17, SD = 15.41, t(794.89) = 2.30, p = 0.02].

**Table 1.** Characteristics of the total sample and the clusters based on chemosensory recovery. Significant differences between the two recovery groups are marked in bold.

| Variable | Full sample<br>(N = 974) | Partial<br>chemosensory<br>recovery (N =<br>471)<br>mean (SD) or N | Substantial<br>chemosensory<br>recovery (N =<br>503)<br>mean (SD) or N | Statistic |
| --- | --- | --- | --- | --- |
| Smell recovery | 41.14 (35.69) | 13.37 (24.97) | 67.16 (21.97) | **t = -35.59,**<br>**p <0.0001** |
| Taste recovery | 40.35 (35.58) | 10.74 (21.58) | 68.07 (20.62) | **t = -42.32,** **$p < 0.0001$** |
| **Chemesthesis recovery** | 28.71 (34.87) | 7.55 (24.08) | 48.52 (31.63) | **$t = -22.83$**<br>**$p < 0.0001$** |
| **Region of residency (Lombardy)** | 653 (67%) | 292 (61.9%) | 361 (71.8%) | $\chi^2 = 2.02$,<br>$p = 0.15$ |
| **Gender (female)** | 675 (69.3%) | 329 (48.7%) | 346 (51.2%) | $\chi^2 = 0.0002$<br>$p = 0.98$ |
| **Age** | 41.33 (11.37),<br>range = 19 - 78 | 43.16 (11.74),<br>range = 19 - 75 | 39.63 (10.75),<br>range = 19 - 78 | **$t = 4.88$,**<br>**$p < 0.0001$** |
| **Onset of symptoms (days)** | 41.61(20),<br>range = 3 - 177 | 43.15 (23.87),<br>range = 3 - 177 | 40.17 (15.41),<br>range = 7 - 152 | **$t = 2.30$,**<br>**$p = 0.02$** |
| **COVID-19 diagnosis** | 591 Self - diagnosed (60.6%),<br>196 Lab tested (20.1%),<br>187 Clinical assessment (19.2%) | 279 Self - diagnosed (59.2%),<br>107 Lab tested (22.7%),<br>85 Clinical assessment (18.02%) | 312 Self - diagnosed (62.02%),<br>89 lab tested (17.7%),<br>102 Clinical assessment (20.3%) | $\chi^2 = 0.81$,<br>$p = 0.66$ |
| **Smokers (yes)** | 427 (43.8%) | 194 (41.18%) | 233 (46.3%) | $\chi^2 = 0.35$,<br>$p = 0.55$ |
| **Prior medical conditions (% based on presence of at least one prior medical condition)** | 311 (31.9%) | 155 (32.9%) | 156 (31.01%) | $\chi^2 = 0.02$<br>$p = 0.89$ |

### Paths from chemosensory loss to recovery

To understand whether partial or substantial recovery from chemosensory loss is dependent on the specific chemosensory loss experienced during COVID-19, we investigated the relationships between clusters of chemosensory loss and recovery. The best clustering profile for chemosensory loss in this dataset resulted to be 3 (bootstrapped stability = 0.93): Cluster 1) moderate smell/taste loss and preserved chemesthesis (N=132; centroids: smell = -20.21, taste = -19.80, chemesthesis= -10.71) ; Cluster 2) substantial smell, taste, and chemesthesis loss (N=516; centroids: smell = -89.4, taste = -90.16, chemesthesis= -76.61); Cluster 3) substantial smell and taste loss, but preserved chemesthesis (N=326; centroids: smell = -87.03, taste = - 65.41, chemesthesis= -14.27).

The majority of individuals with moderate smell/taste loss and preserved chemesthesis (χ^2^ (2)= 26.92, p < 0.001; post-hoc p < 0.001) reported a partial recovery (24.8%, N = 117), and only the 2.9% (N = 15) reported substantial recovery. On the contrary, most of the individuals with substantial loss of smell, taste, and chemesthesis showed the highest rate of recovery (67.6%, N = 340; partial recovery: 37.4%, N = 176; post-hoc p < 0.001). Among the individuals who reported substantial smell and taste loss, but preserved chemesthesis there was no significant difference in the reported recovery (substantial recovery: 29.4%, N = 148; partial recovery: 37.8%, N = 178; post-hoc p = 0.38; see Figure 3). Noteworthy, the clusters “moderate smell/taste loss and preserved chemesthesis” and “substantial smell and taste loss, but preserved chemesthesis” together account for the 62% of the partial recovery cluster, while only for the 32% of the substantial one (χ2 (2)= 46, p < 0.001).

**Figure 3.**
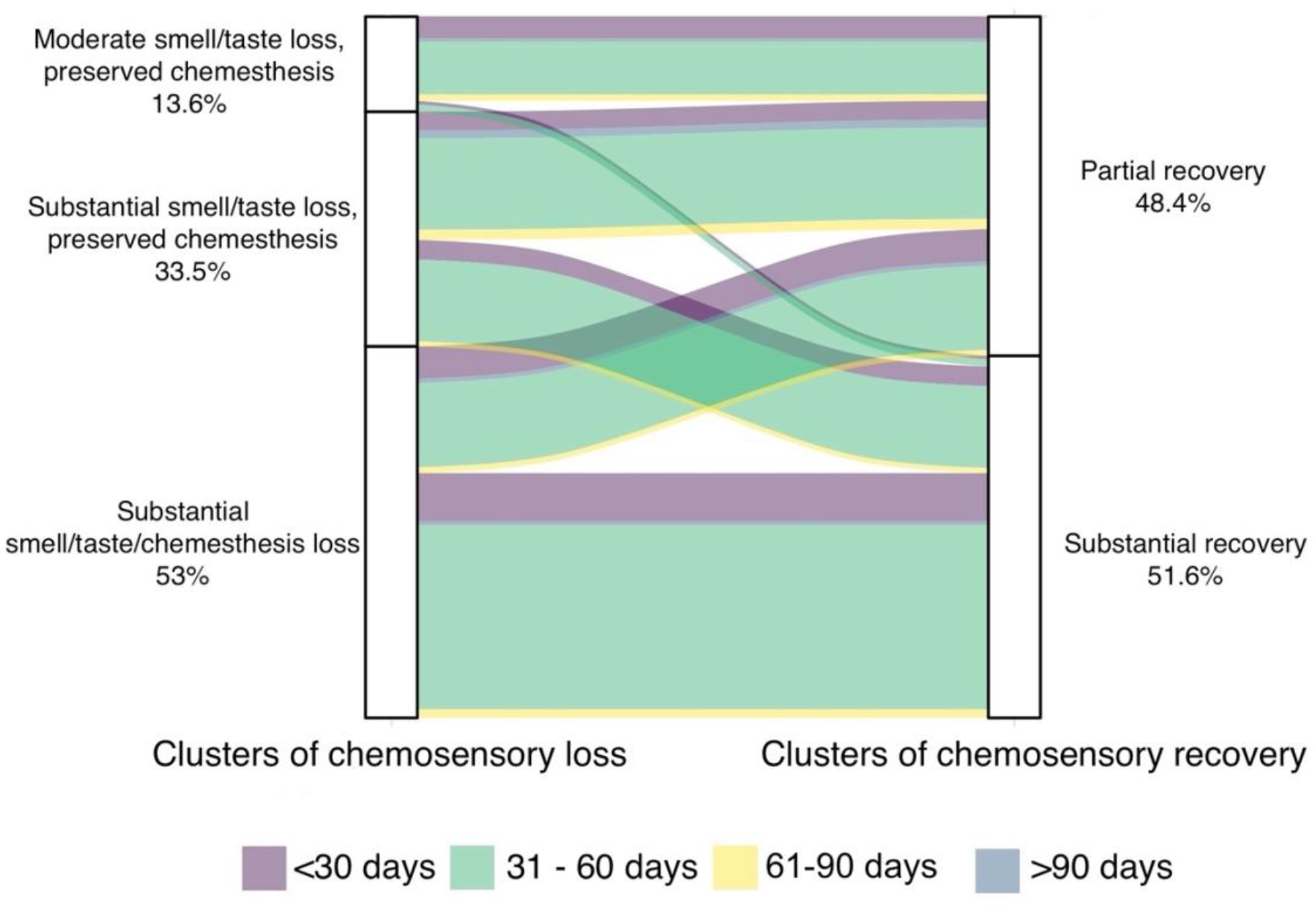
The pattern of chemosensory loss and recovery clusters in relation to days between the date of onset and completion of the questionnaire.

Most participants (71.45%, N = 696) reported the onset of the symptoms within a 31-60 days time frame before the completion of the questionnaire. There was no significant difference in the distribution of the chemosensory recovery groups or the chemosensory loss groups on the different time frames of the onset of symptoms (recovery groups: χ^2^ (3)= 3.35, p = 0.34; loss groups: χ^2^ (6)= 4.87, p = 0.56; see Figure 3).

### Association of chemosensory recovery with demographic and clinical predictors

To account for individual differences in baseline chemosensory abilities, and in the use of rating scales, we use as indicators of the status of the chemosensory functions, the “recovery” as the difference between ratings of patients’ chemosensory abilities after- and during- the respiratory illness and the “loss’’ as the difference between ratings of their chemosensory abilities during- and before- the respiratory illness (Table 2; see the “Methods” section). The model on smell recovery showed a significant main effect of *regions* (Lombardy, Other Regions), indicating that participants living in Lombardy reported higher levels of smell recovery (mean = 42.90, sd = 35.90) compared to participants living in other regions (mean = 37.58, sd = 35.04); a significant main effect of *age*, indicating that younger participants reported higher smell recovery; and a significant main effect of *index of smell loss* (During - Before ratings), indicating higher recovery when participants reported greater smell loss (see Figure 2s of the Supplemental material for the visualization of main effects). Lastly, there was a significant interaction between *regions* (Lombardy, Other Regions) and *time from onset* (number of days from the reported date of the symptoms onset of respiratory illness, and the date of survey completion; Figure 4A). Post-hoc tests showed that in participants residing in Lombardy, when the time from the onset of the disease was longer this was associated with lower smell recovery [t(1) = -3.57, p < 0.001]; this effect is not present in participants of other Italian regions [t(1) = -0.17, p = 0.86] and could be related with the high incidence and the correlated logistic problem of the health system that Lombardy experienced during the first wave. Indeed, Lombardy registered one-third of all cases and half of all deaths in Italy^49,58^.

**Table 2.** Summary of the linear regressions on smell, taste, and chemesthesis recovery

| Smell Recovery | Estimate | SE | t value | Pr(>\|t\|) |
| --- | --- | --- | --- | --- |
| (Intercept) | 19.720 | 8.545 | 2.308 | **0.0212** |
| Region (*Lombardy*) | 14.222 | 5.096 | 2.791 | **0.0054** |
| Type of diagnosis (*Clinical assessment*) | -20.783 | 9.814 | -2.118 | **0.0345** |
| Type of diagnosis (*Self-diagnosed*) | -9.660 | 7.491 | -1.290 | 0.1975 |
| Number of symptoms | -1.438 | 0.814 | -1.766 | 0.0776 |
| Time from onset | -0.013 | 0.078 | -0.172 | 0.8635 |
| Smoking (*Yes*) | 3.180 | 2.125 | 1.497 | 0.1348 |
| Index of smell loss | -0.462 | 0.035 | -13.115 | **<0.001** |
| Age | -0.226 | 0.093 | -2.419 | **0.0158** |
| Region (*Lombardy*): Time from onset | -0.242 | 0.106 | -2.276 | **0.0231** |
| Type of diagnosis (*Clinical assessment*):<br>Number of symptoms | 2.541 | 1.181 | 2.152 | **0.0316** |
| Type of diagnosis (*Self-diagnosed*): Number of symptoms | 1.614 | 0.964 | 1.674 | 0.0944 |
| Residual standard error: 32.53 on 962 degrees of freedom |  |  |  |  |
| Multiple R-squared: 0.1787, Adjusted R-squared: 0.1693 |  |  |  |  |
| F-statistic: 19.03 on 962 and 11 DF, p-value: <0.001 |  |  |  |  |
| **Taste Recovery** | **Estimate** | **SE** | **t value** | **Pr(>\|t\|)** |
| (Intercept) | 8.514 | 5.395 | 1.578 | 0.1149 |
| Region (*Lombardy*) | 15.249 | 4.565 | 3.340 | **0.0009** |
| Time from onset | 0.063 | 0.071 | 0.887 | 0.3756 |
| Smoking (*Yes*) | 4.060 | 1.934 | 2.099 | **0.0361** |
| Prior conditions | 1.999 | 1.278 | 1.564 | 0.1182 |
| Index of taste loss | -0.564 | 0.030 | -18.756 | **<0.001** |
| Age | -0.411 | 0.086 | -4.759 | **<0.001** |
| Region (*Lombardy*): Time from onset | -0.271 | 0.097 | -2.797 | **0.0053** |
| Residual standard error: 29.73 on 966 degrees of freedom |  |  |  |  |
| Multiple R-squared: 0.3071, Adjusted R-squared: 0.302 |  |  |  |  |
| F-statistic: 61.15 on 966 and 7 DF, p-value: <0.001 |  |  |  |  |
| **Chemesthesis Recovery** | **Estimate** | **SE** | **t value** | **Pr(>\|t\|)** |
| (Intercept) | 12.336 | 5.826 | 2.117 | **0.0345** |
| Type of diagnosis (*Clinical assessment*) | -14.728 | 7.409 | -1.988 | **0.0471** |
| Type of diagnosis (*Self-diagnosed*) | 0.490 | 5.633 | 0.087 | 0.9307 |
| Number of symptoms | -0.926 | 0.616 | -1.503 | 0.1330 |
| Smoking (*Yes*) | 3.672 | 1.605 | 2.288 | **0.0223** |
| Index of chemesthesis loss | -0.650 | 0.021 | -30.676 | **<0.001** |
| Age | -0.282 | 0.070 | -4.026 | **0.0001** |
| Type of diagnosis (*Clinical assessment*):<br>Number of symptoms | 2.300 | 0.895 | 2.571 | **0.0103** |
| Type of diagnosis (*Self-diagnosed*): Number of symptoms | 0.374 | 0.729 | 0.512 | 0.6085 |
| Residual standard error: 24.66 on 965 degrees of freedom |  |  |  |  |
| Multiple R-squared: 0.5038, Adjusted R-squared: 0.4997 |  |  |  |  |
| F-statistic: 122.5 on 965 and 8 DF, p-value: <0.001 |  |  |  |  |

Note: Contrast condition from the reference for categorical factors is reported in italic inside brackets. Significant differences are marked in bold. SE = standard error. Table shows models with *Other regions* (Region variable), *Lab tested* (Type of diagnosis variable), *No* (Smoking variable) set as references.

**Figure 4.**
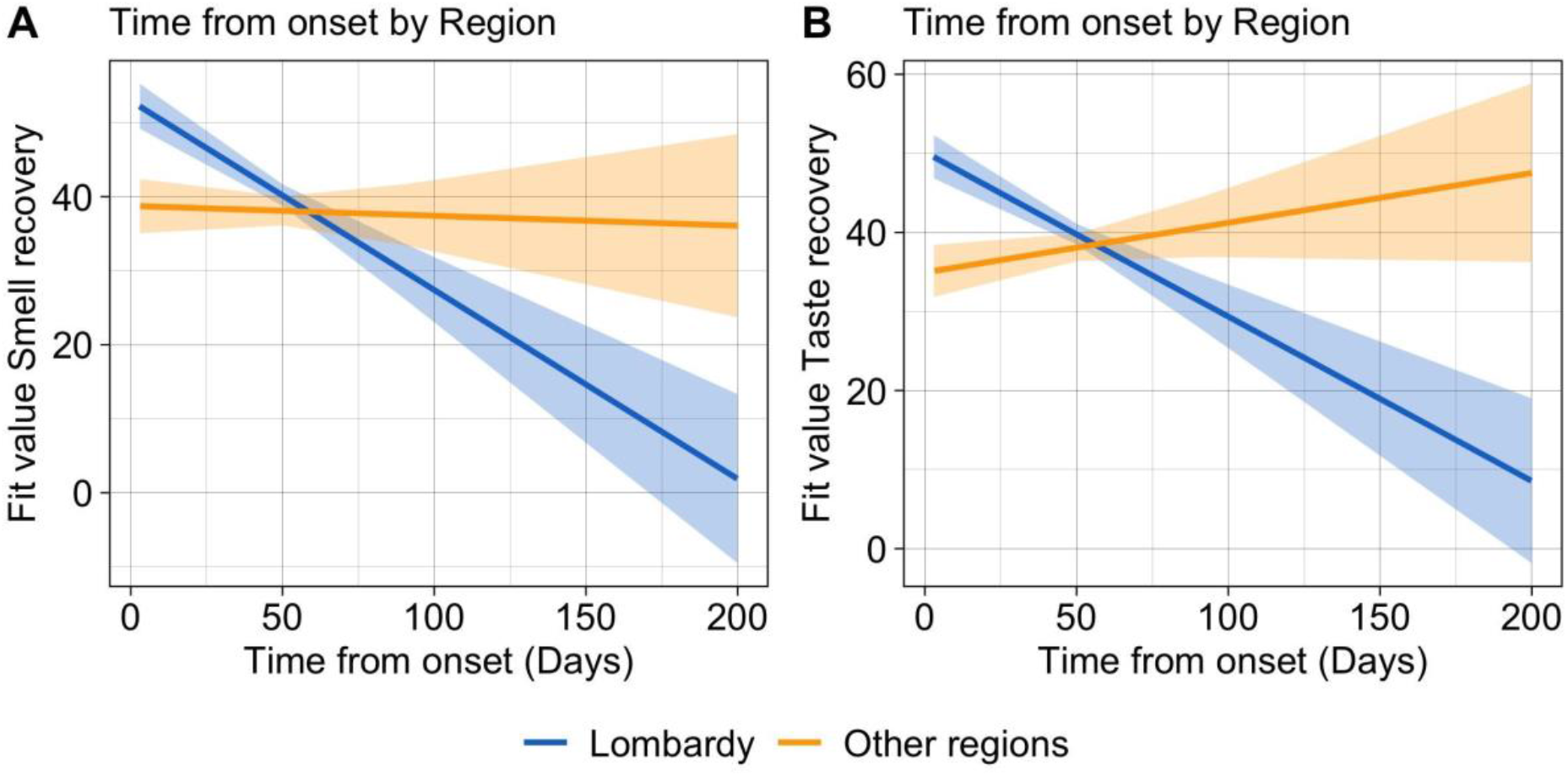
Representation of significant interaction effects of the regression models. Fitted lines of the time from onset and region interaction effects on A) smell recovery and B) taste recovery.

The model on taste recovery showed a main effect of *age*, indicating that older participants reported less taste recovery and a significant main effect of *taste loss* (During - Before ratings), consistent with higher recovery when participants reported higher taste loss. Effect of *Smoking* (yes, no) was significant as well, suggesting that smokers reported higher taste recovery (mean = 43.04, sd = 34.98) compared to non-smokers (mean = 38.24, sd = 35.93; Figure 3s in the Supplemental material for visualization of main effects). This observation is controversial^59,60^, as recent data suggest, smokers risk a more severe course of the disease^59,61,62^. However, we cannot exclude that this effect could be the result of the temporary abstinence from smoking during the disease as it has been reported that the effects of smoking on chemosensory function are short-term^22^. Moreover, a significant interaction between *regions* and *time from onset* (Figure 4B) was observed. Post-hoc analyses showed that a longer time from onset of respiratory symptoms was associated with lower taste recovery only in participants from the Lombardy region [t(1) = -3.19, p = 0.0014]. Moreover, data show that differences in taste recovery among regions are greater [t(1) = 3.19, p < 0.001] in participants with longer time from onset, than in participants with shorter time from onset [t(1) = 3.25, p = 0.0012].

Results on chemesthesis recovery indicate a significant main effect of *age*, showing that older participants reported a smaller index of recovery than younger participants; a significant main effect of *chemesthesis loss* (During - Before ratings) showed high recovery when it was reported as more pronounced; and a significant main effect of *smoking*, showing that smokers reported higher level of chemesthesis recovery (Figure 4s in the Supplemental material for visualization of main effects). Post-hoc analyses were used to resolve the interaction between *the number of symptoms* (as the total sum of the reported symptoms experienced with the respiratory illness) and *type of diagnosis* (self-diagnosed, clinical assessment, lab tested). Post - hoc contrasts show that participants who received a respiratory illness diagnosis after a clinical assessment [t(1) = 2.11, p = 0.034] had a greater level of chemesthesis recovery despite the elevated number of symptoms. This effect was not significant in participants whose diagnosis was determined by lab test [t(1) = -1.50, p = 0.13] or by self-diagnosed (Figure 5) groups.

**Figure 5.**
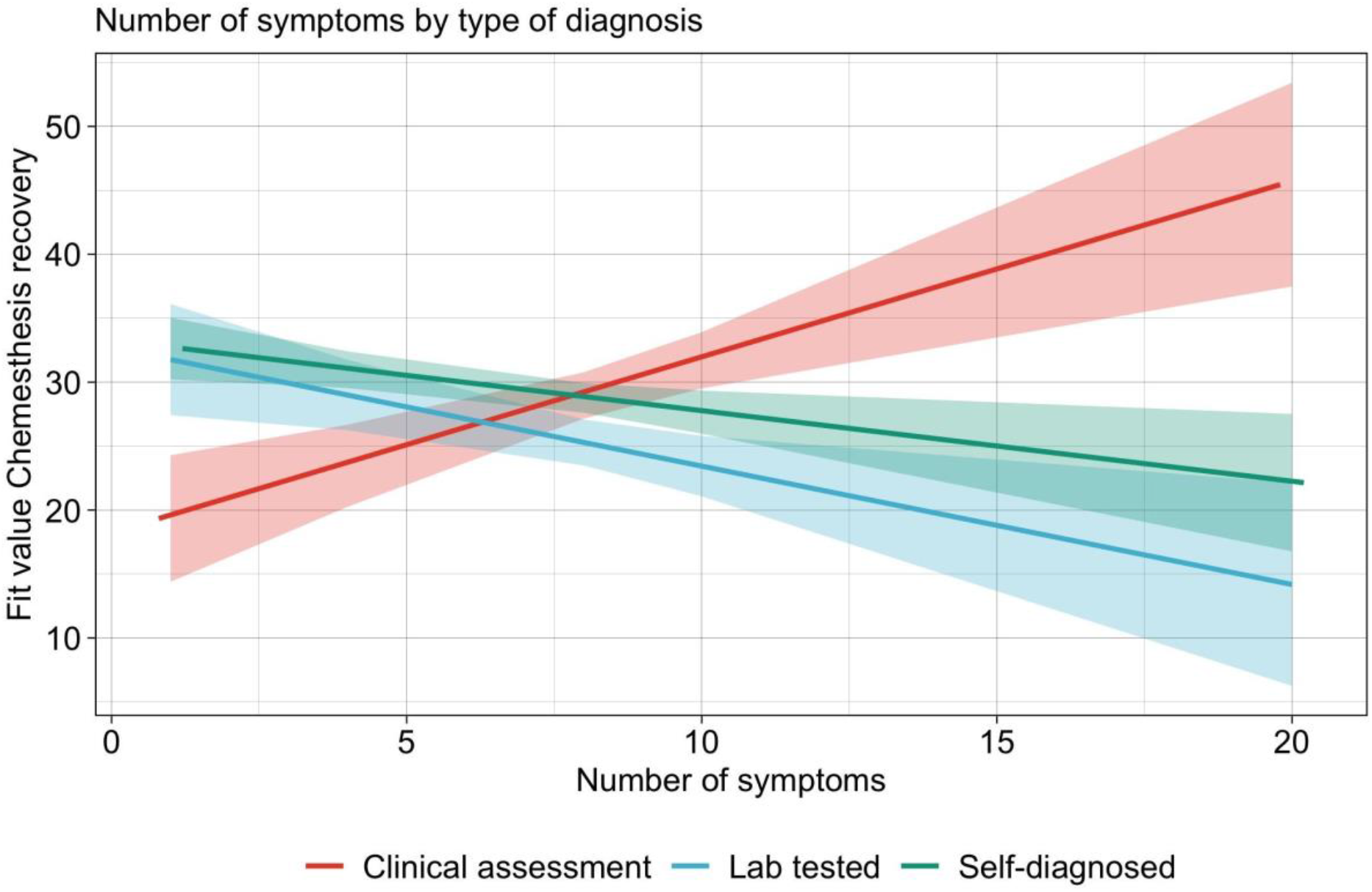
Representation of significant interaction effects of the regression models. Fitted lines of the number of symptoms and type of diagnosis on chemesthesis recovery.

## Discussion

Since the beginning of the COVID-19 pandemic, an increased number of patients with taste and smell loss has been reported, and an increased evidence emerged in the literature reporting chemosensory deficits as a salient feature of the disease^1^. The present study aimed to characterize on a larger scale the persistence and recovery process of chemosensory deficits associated with SARS-CoV-2 infection, attempting to delineate the expectations of recovery for the patients as well as predict/identify groups of patients in need of additional post-viral care.

Despite some limitations^24,57^, the analysis of self-reports of patients’ chemosensory abilities is to date the most effective strategy to target the largest number of patients that could not be otherwise reached because of the safety policies implemented during the COVID-19 pandemic, as well as the lack of widespread routine chemosensory testing. Current standardized evaluations of smell and taste for clinical purposes require lengthy and maskless testing sessions, thus making this practice unsafe^63,64^. Additionally, even if this practice would be safe, it is not common knowledge among first-line healthcare professionals. Therefore, in order to recognize early during the disease and characterize over time these extremely common long-term consequences of COVID-19, it is of paramount importance to add to the first patients’ assessment a set of well-framed informed questions on smell and taste loss and recovery. A direct comparison of the objective and subjective chemosensory assessment showed that subjective methods (self-reports) might underestimate chemosensory loss in COVID-19 patients^1^, nevertheless, self-reports can provide a first-aid tool to estimate chemosensory loss among the population, with a significant impact on the patients’ care and quality of life. A first indication emerging from our analysis is that asking the patient to rate their smell, taste, and ability to perceive chemical irritation (chemesthesis) on a 0-100 visual analog scale (VAS) before, during, and after the resolution of the respiratory symptoms is a first and important step to understand the recovery. Importantly, chemesthesis, primarily mediated by the trigeminal nerves, is not only responsible for the detection of chemical irritants but it is also involved in inflammatory responses. Most recent reports suggest that this inflammatory response is a possible contributor to the exacerbation of the tissue damage induced by viral Sars-CoV 2 infection^65,66^. In patients experiencing chemesthesis loss, inflammatory responses might be reduced or impaired, decreasing the probability of further damage to the olfactory epithelium. Interestingly, 62.2% of the subjects experiencing partial chemosensory recovery showed no chemesthesis loss (37.4% moderate smell/taste loss, and 24.8% substantial smell/taste loss), supporting the hypothesis of a contribution of the inflammatory response to long term chemosensory loss. From these observations, it emerges that the evaluation of chemesthesis function, which has been mostly neglected outside of the GCCR efforts (Parma et al. Gerkin et al.), might provide healthcare professionals not only with an indication of the outcome of the chemosensory recovery process, but also inform the design of better strategies for early treatment of post-viral symptoms.

To account for individual differences in baseline chemosensory abilities, and in the use of rating scales, we suggest to use as indicators of the status of the chemosensory functions, the “recovery” as the difference between ratings of patients’ chemosensory abilities after - and during- the respiratory illness and the “loss’’ as the difference between ratings of their chemosensory abilities during - and before- the respiratory illness. By using olfactory, taste and chemesthesis loss as indicators, we can predict the level of the recovery of each chemosensory ability within the time frame of 120 days. Indeed, our data show that greater sensory loss corresponds to a greater rate of recovery. Although it has been shown that subjective ratings are a good proxy for the understanding of chemosensory loss during the COVID-19 pandemic^50,57^ (and summarized by^67^) especially in relation to complete and sudden smell loss, these measurements might suffer from underreporting and over-reporting biases^68,69^ and possible arbitrary scale usage. Participants who experienced a more severe chemosensory loss might tend to overestimate their recovery. Other studies which used objective testing methods, also observed a similar dependency between loss and recovery, strengthening the evidence that a greater olfactory improvement post-infection is more likely in patients experiencing sudden anosmia or ageusia during the viral infection than in those experiencing hyposmia and hypogeusia^27,29^.

As observed previously^3,28–30,32,70,71^, a demographic factor that should be considered is age. We performed a first exploratory cluster analysis (Figure 2A) that suggested two different patterns of chemosensory recovery: one is characterized by moderate smell, taste, and chemesthesis recovery; and a second one by a substantial smell, taste, and chemesthesis recovery. These two clusters significantly differ for age of the subjects, with the first group on average older than the second. Our analysis then confirms the role of age in the recovery from the chemosensory deficits, showing that younger participants are associated with a better chemosensory recovery prognosis than older ones for all three chemosensory modalities. Although age-related differences in chemosensory abilities are well known, in the case of COVID-19 this relationship is less clear and still controversial. Results from several studies^28,32,71^ did not find any age-related difference, while Moein et al.^5^, with an analytic approach similar to ours, found that older age had a negative impact on smell recovery, which is in agreement with our results on this dataset.

Another interesting aspect of our analysis is that smokers report greater recovery rates for taste and chemesthesis than non-smokers. We cannot exclude that this effect could be the result of the temporary abstinence from smoking during the disease or the overall limited severity of the disease of participants responding to a survey online. It has been reported that the effects of smoking on chemosensory function are short-term^22^. While being a smoker could thus be a confounding question to ask, its statistical link with taste and chemesthesis recovery could improve the prognosis.

Finally, our analysis of demographic and clinical predictors for recovery of each sensory modality reveals that being resident in Lombardy was predictive of greater smell recovery. Indeed, Lombardy was the epicenter of the first wave of the COVID-19 pandemic in Italy, with an overall earlier date of onset and the registering of the highest number of cases (and survey participants) in comparison to the rest of Italy. Differences that emerged between Lombardy and other regions could be due to differences in the regional management of the pandemic, but also to the delayed spreading of the disease in the other regions which registered a relatively low number and later onset cases in comparison to Lombardy in the time frame we analyzed^49,58^. In Lombardy, the time of onset of the disease is predictive of a worst prognosis of chemosensory recovery, confirming the presence of a group of patients whose recovery from any of the symptoms does not occur within 4-6 weeks from their onsets^72^. This group deserves further investigation, given that the sequela from non-COVID-19 post-viral chemosensory loss can last on average 1 years^73^. While regional differences that emerged from our analysis could not be used as a first-aid tool to understand the recovery directly by local healthcare professionals, they could help in understanding the epidemiological scenario of the pandemic.

## Conclusions

With the SARS-CoV-2 pandemic, the number of patients affected by chemosensory loss substantially increased. Our work provides indications on the recovery process on which we shaped a scientific-based approach for healthcare professionals to characterize the clinical picture of patients reporting chemosensory loss due to COVID-19 infection. We further provide indexes such as loss and recovery that would be extremely useful for single ENT doctors to have a starting point for further diagnosis and prognosis. Three different profiles of chemosensory loss were identified: substantial loss of all the three chemosensory modalities, substantial loss of only smell and taste, and moderate loss of only smell and taste. Clinicians must take into account demographic factors that influence chemosensory recovery, among them the age as we showed that older adults had a longer recovery period. Uncovering the self - reported phenomenology of recovery from smell, taste, and chemesthetic disorders is the first, yet essential step, to provide healthcare professionals with the tools to take purposeful and targeted action to address chemosensory disorders and its severe discomfort.

## Method

### The GCCR online survey

The data utilized in this study is part of the GCCR survey^57^, which was developed as a global, crowdsourced online study, and deployed in 35 languages. The survey aimed to measure self - reported smell, taste, and chemesthesis function, and nasal blockage, amongst other variables, in participants with recent (within the past two weeks) or current respiratory illness, including COVID-19. Participants were asked to rate their ability to smell, taste, and perceive cooling, tingling, and burning sensations (chemesthesis) before, during, and, in case of recovery, after their respiratory illness, using 100-point visual analog scales (VAS). The online survey was approved by the Office of Research Protections of The Pennsylvania State University (STUDY00014904) in accordance with the revised Declaration of Helsinki.

### Participants

The entry criterion for participation in the GCCR survey was a recent or current respiratory illness (symptoms present in the past 2 weeks). Accordingly, only participants who answered “Yes” to Question 6, “Within the past two weeks, have you been diagnosed with or suspect that you have a respiratory illness?” were allowed to complete the survey (see Appendix 1 of Parma et al.^57^ for all survey questions). In the present study, were included only participants who reported to be resident in Italy (n = 5564) and a COVID-19 diagnosis or symptoms [Question 8 “Have you been diagnosed with COVID-19?”, answers “No-I was not diagnosed, but I have symptoms” (self-diagnosed group), “Yes-diagnosed based on symptoms only” (Clinical assessment group), “Yes-diagnosed with viral swab”, “Yes-diagnosed with another lab test”, (Lab tested group)] (n = 1647). In order to investigate chemosensory recovery, we included only participants who answered “Yes - partly” or “Yes - fully” to Question 28 “Have you recovered from your recent respiratory Illness or diagnosis?” (n = 1335). Other exclusion criteria were: incomplete ratings (n=167), no date of onset of respiratory illness symptoms provided (n = 166; Question 7: “What date did you first notice symptoms of your recent respiratory illness?”), inconsistent responses in questions on smell changes (n = 22; specifically, selecting changes in smell in Question 10 “Have you had any of the following symptoms with your recent respiratory illness or diagnosis?”, reporting a difference in Question 13 “Rate your ability to smell before your recent respiratory illness or diagnosis” and/or select at least one answer from Question 15 “Have you experienced any of the following changes in smell with your recent respiratory illness diagnosis?”), age above 100 (n = 1), reported date of onset of respiratory symptoms after the date of participation or before January 2020 (n=5). The final sample included 974 participants.

### Indices

To standardize statistical analyses, some measures were combined into indices. We defined the *time from onset* as the number of days from the reported date of symptoms onset of respiratory illness and the date of survey completion. We defined the *number of symptoms* as the total sum of the reported symptoms experienced with the respiratory illness (“Have you had any of the following symptoms with your recent respiratory illness or diagnosis?” Question 10) and the *prior conditions* as the total sum of the reported medical conditions experienced in the 6 months prior the onset of the respiratory illness (“Did you have any of the following in the 6 months prior to your recent respiratory illness or diagnosis?” Question 38). Moreover, *indices of loss* for smell, taste, and chemesthesis was computed by subtracting ratings “before illness” (Question 14 “Rate your ability to smell BEFORE your recent respiratory illness or diagnosis”) from ratings “during illness” (Question 13 “Rate your ability to smell DURING your recent respiratory illness or diagnosis”). Finally, *indices of recovery* of each sense (Smell, taste, and chemesthesis) were computed by subtracting ratings “during illness” from ratings “after illness” (Question 29 “Rate your ability to smell AFTER your recovery”).

### Statistical analyses

Data were pre-processed and analyzed using the software R^74^. Statistical analyses were pre - registered at the Open Science Framework (OSF, https://osf.io/vun72/) before the data became available. First, to investigate whether chemosensory profiles of recovery exist and if they followed the profiles found for chemosensory loss^57^, we extended the cluster analysis of Parma et al.^57^ on the Italian dataset, that only partially overlapped with the data previously analyzed (594 Italian residents^57^). Cluster analyses were performed based on the similarities and differences in indexes of smell, taste, and chemesthesis loss, and recovery using the *k-means* function from the R default stats package. The optimal number of clusters was determined with *NbCluster*^75^, which tests 30 methods that vary the combinations of cluster numbers and distance measures for the *k-means* clustering. Cluster stability was estimated through a bootstrapping approach (100 iterations) with the *bootcluster* package^76^. Descriptive analyses on the resulting clusters were run using t-tests (stats package^77^) and chi-square tests (chisq.test function of the stats package^78^). Next, smell, taste, and chemesthesis recovery were investigated through three separate multiple linear regression models (*lm* function of *stats* package) with the same predictors. Predictors included continuous and categorical variables. The former included: number of symptoms, time from onset, prior conditions, and index of the chemosensory loss related to the dependent variable (e.g. smell loss for smell recovery); the latter included: region of residence (Lombardy, Other regions), type of diagnosis (Self - diagnosed, Clinical assessment, or lab tested), smoking (yes, no; also including e-cigarette). In order to explore the recovery profile and region specificity, in the models, we included interaction between these variables: region of residence, type of diagnosis, number of symptoms, and time from onset. To avoid overly complicated and uninterpretable models, only second-level interactions were included. To ensure that each predictor improved the models’ fit, the function *step* (*stats* package) was used to perform automatic backward elimination, which relies on the AIC criterion^79^. Factors that did not significantly improve the models’ fit were removed. AIC values of the initial and final models were calculated using the ANOVA function (stats package^77^). Collinearity was calculated with the Variance Inflation Factors (VIF) using the *vif* function of the car package^80^. Interactions including continuous factors were analyzed according to Aiken & West’s method^81^.

In the light of recent studies from the GCCR dataset^24,57,82^, additions to the pre-registered linear models were necessary: 1) smell, taste and chemesthesis ratings were not analyzed as repeated measures (before, during, after) but rather indices of loss and recovery were computed to better characterized the degree of changes; 2) since in cluster analyses age was significantly different between the two clusters, it was included as fixed and not anymore as a random factor; 3) gender and type of recovery were removed because they did not improve the models’ fit. Due to the particular spread of the pandemic in Italy, the region of residence was also included as a predictor.

## Data Availability

The preprocessed datasets and the R code for the reported analyses can be found on the Open Science Framework database (https://osf.io/vun72/)

