## Supplemental figures for "From loss to recovery: how to effectively assess chemosensory impairments during COVID-19 pandemic"

**Supplementary material**

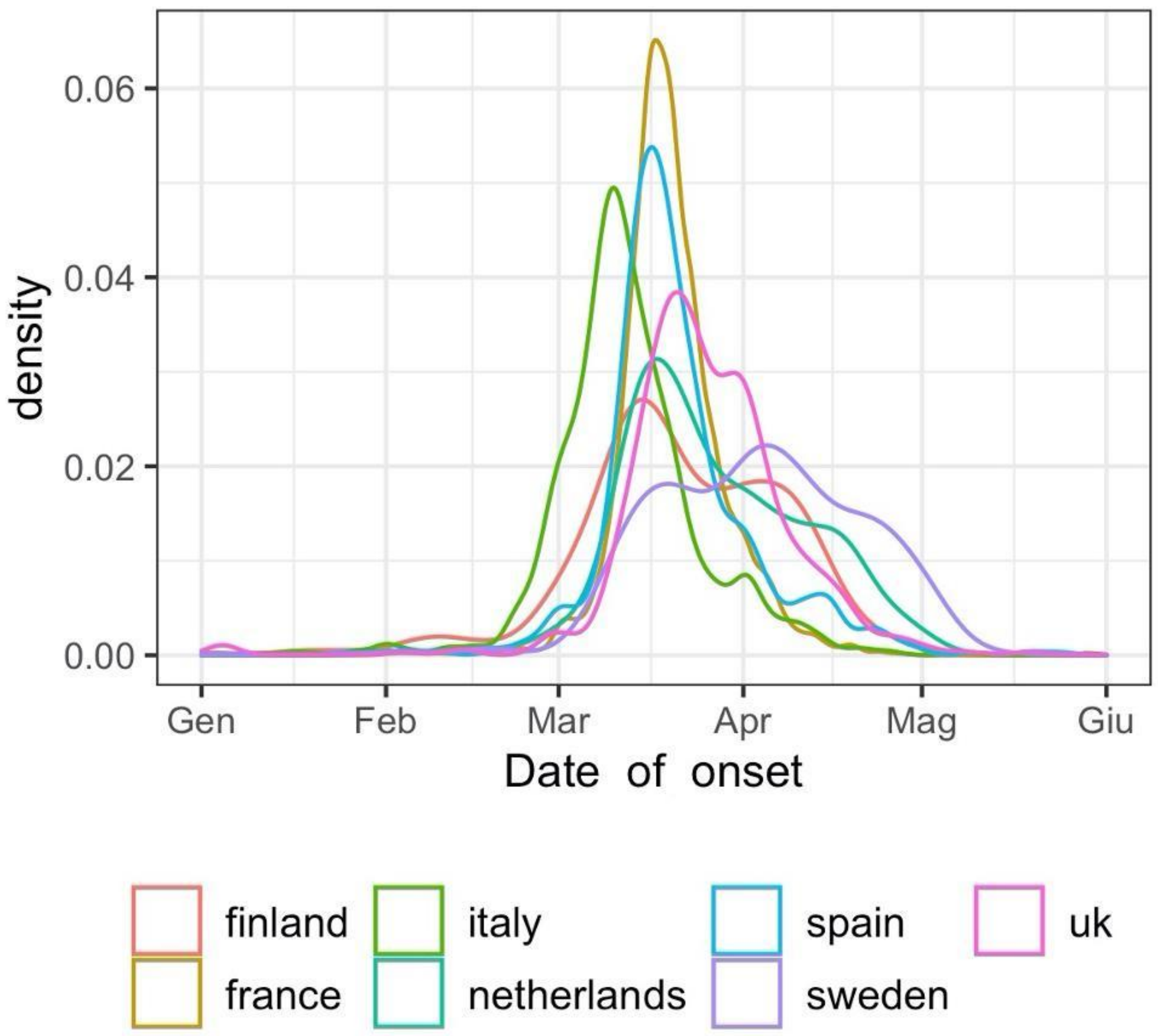


**Figure 1s.** Dates of COVID-19 onset for different countries (A). In agreement with other epidemiological data Italy is the first country to be hit by the disease (green curve) followed by Spain (cyan) and France (gold).

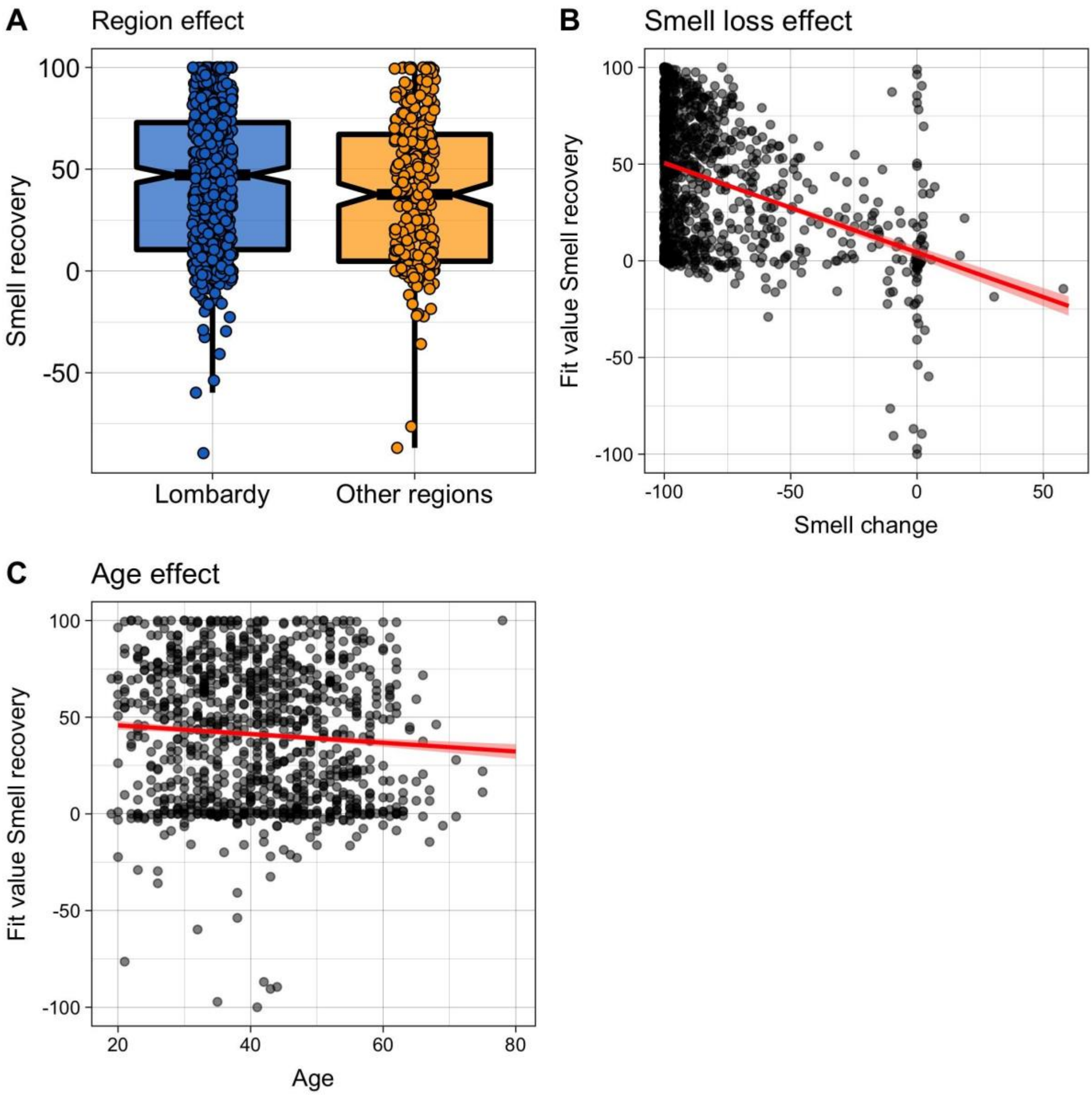


**Figure 2s.** Significant main effect predictors of smell recovery. A) Data distribution by region. Boxplots depict the median (horizontal black line) and quartile ranges of the distribution, whiskers indicate maximum and minimum values, colored dots represent data distribution. B) and C) Main effect of age and smell loss, with data distribution as black and grey dots and fitted lines in red.

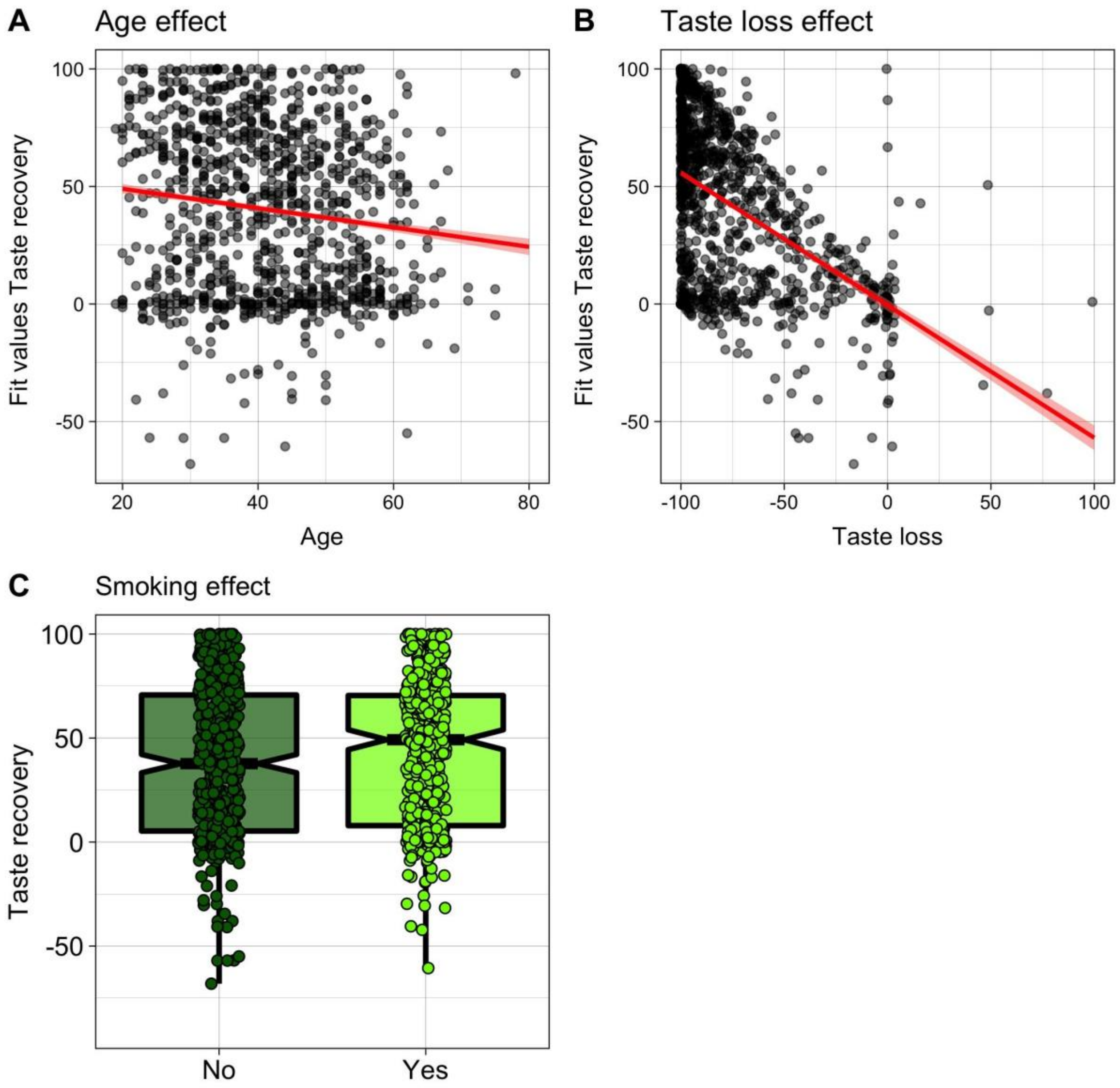


**Figure 3s.** Significant main effect predictors of taste recovery. A) and B) Main effect of age and taste loss, with data distribution as black and grey dots and fitted lines in red. C) Data distribution by smoking. Boxplots depict the median (horizontal black line) and quartile ranges of the distribution, whiskers indicate maximum and minimum values, colored dots represent data distribution.

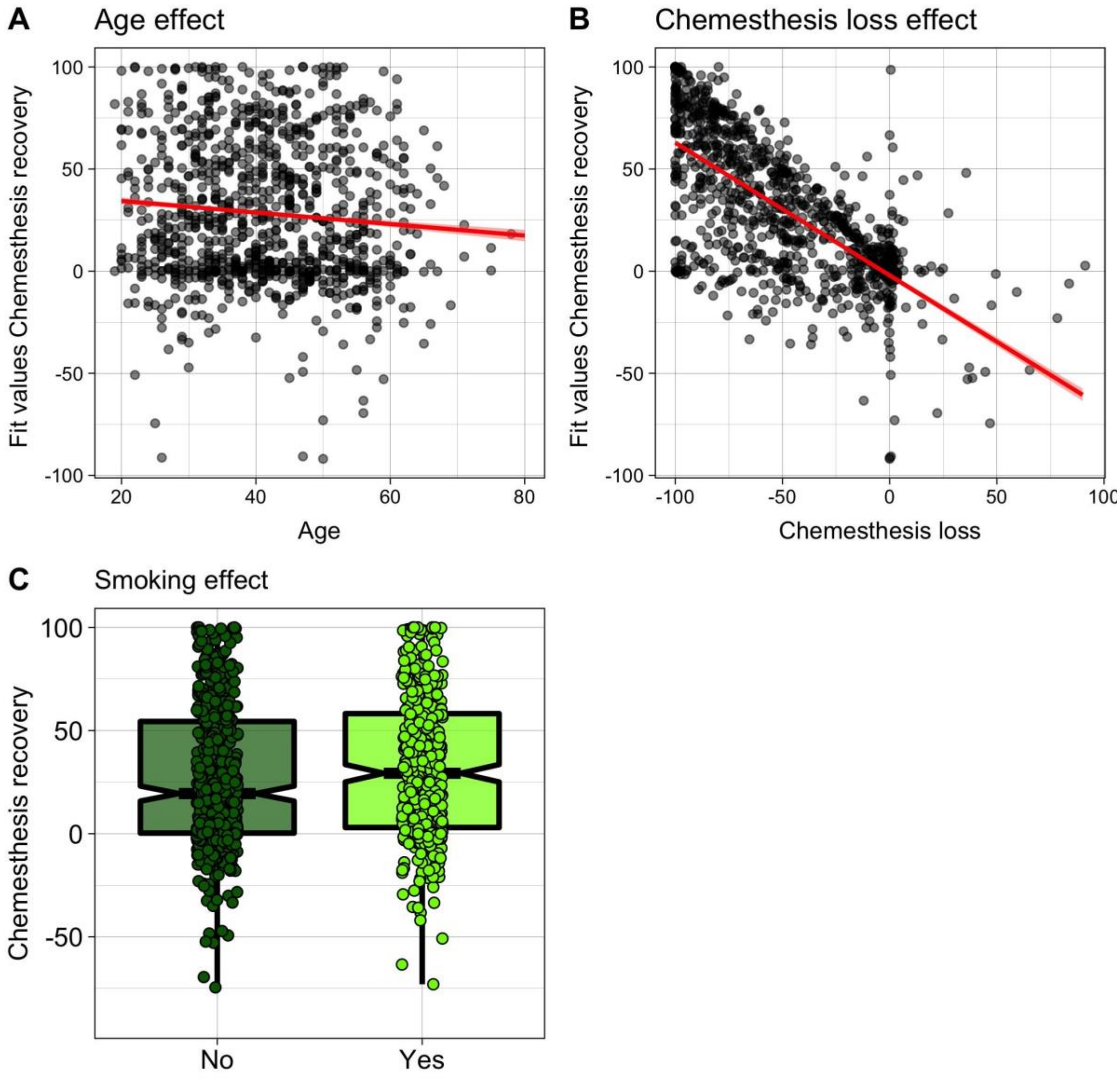


**Figure 4s.** Significant main effect predictors of chemesthesis recovery. A) and B) Main effect of age and chemesthesis loss, with data distribution as black and grey dots and fitted lines in red. C) Data distribution by smoking. Boxplots depict the median (horizontal black line) and quartile ranges of the distribution, whiskers indicate maximum and minimum values, colored dots represent data distribution.
